# Accurate machine learning-based CVD risk prediction in primary care may reduce the need for routine health care checks

**DOI:** 10.1101/2025.06.09.25329273

**Authors:** Katarzyna Dziopa, Sophie V Eastwood, Daniel Bos, Maryam Kavousi, Maarten J G Leening, Joline W J Beulens, Peter P Harms, Nishi Chaturvedi, Folkert W Asselbergs, Amand F Schmidt

**Affiliations:** Department of Cardiology, Amsterdam Cardiovascular Science, Amsterdam University Medical Center, University of Amsterdam, Amsterdam, The Netherlands; Institute of Cardiovascular Science, Faculty of Population Health Sciences, University College London, London, United Kingdom; UCL BHF Research Accelerator Centre, London, UK; Departments of Cardiology, Epidemiology, and Radiology, Erasmus MC – University Medical Center Rotterdam, Rotterdam, the Netherlands; Department of Clinical Epidemiology, Harvard School of Public Health, Boston, USA; Department of Epidemiology & Data Science, Amsterdam University Medical Center, Vrije Universiteit Amsterdam, Amsterdam, The Netherlands; Amsterdam Public Health Research Institute, Health Behaviors & Chronic Diseases and Personalized Medicine, Amsterdam, The Netherlands; Department of General Practice Medicine, Amsterdam University Medical Center, Vrije Universiteit Amsterdam, Amsterdam, The Netherlands; Institute of Health Informatics, University College London, London, United Kingdom; The National Institute for Health Research UCL Hospitals Biomedical Research Centre, University College London, London, United Kingdom; Department of Cardiology, University Medical Center Utrecht, Utrecht, The Netherlands

**Keywords:** Missing data, machine learning, Cardiovascular disease, Prediction, Risk Score

## Abstract

**Background:** Cardiovascular risk prediction models, such as PCE, QRISK3, and SCORE2 are recommended tools to guide treatment initiation/intensification in primary care. In clinical practice, the absence of one or more required predictors is common, which precludes routine application of such models.

**Methods:** We developed a set of partial models predicting the 10-year risk of cardiovascular disease (CVD) and major CVD (additionally considering atrial fibrillation, heart failure, and peripheral arterial disease) using combinations of 14 predictors, allowing application in settings were only a subset of variables is available. The set of partial models was evaluated across five studies jointly comprising 105,550 participants.

**Findings:** We trained 4,096 unique models to predict 10-year major CVD risk, observing near identical performance evaluated against CVD and major CVD. The c-statistic ranged between: quartiles (Q1) 0.71 and Q3: 0.73 across the five studies. This was comparable to the performance of the PCE (Q1: 0.70, Q3: 0.74, 10 predictors) and SCORE2 (Q1: 0.71, Q3: 0.75, 8 predictors). Due to large number of required predictors (22/23 for men/women) the QRISK3 was evaluated in a single cohort: c-statistic 0.72 (95% CI 0.72; 0.73). Model performance remained adequate when focussing on the set of partial models using 2-4 predictors: c-statistic Q1: 0.70 and Q3: 0.71. Partial models demonstrated reasonable calibration across most studies, observing a limited risk underestimation in two cohorts. Partial models excluding blood pressure and lipids demonstrated similar performance to models incorporating these variables. The set of partial models has been made available through a python-based application programming interface.

**Interpretation:** We show that in the presence of partially missing data, clinically relevant predictions of the 10-years risk of major CVD can be obtained by using a subset of features, facilitating improved and more timely treatment decisions.

**Funding:** Dutch Research Council, British Heart Foundation, UK Research and Innovation.

**RESEARCH IN CONTEXT:** *Evidence before this study:* Before submitting our article on May 5, 2025, we searched PubMed articles published from database inception, using the terms “missing data” [tiab] or “incomplete data”[tiab], “cardiovascular disease” [tiab], and “risk score” [tiab] or “prediction”[tiab]. Studies unrelated to cardiovascular disease (CVD) prediction were excluded. None of the identified CVD prediction models allowed for missing input data and instead considered missing data solely at the stage of model derivation.

*Added value of this study:* The applicability of widely recommended cardiovascular risk prediction models, such as SCORE2 (Europe), PCE (US), and QRISK3 (UK), is constrained by the need to measure all included variables. The absence of even a single variable, such as total cholesterol used in all three aforementioned models - precludes risk prediction. For instance, among individuals aged 40 to 69 years without a history of cardiovascular disease, only 10.8% have a recorded cholesterol measurement at any point in their medical history. To overcome these limitations, this study introduces an approach using 4,096 partial models to predict 10-year risk of (major) cardiovascular disease using combinations of 14 variables, specifically designed to address the challenge of missing data. Performance was assessed across five datasets from the UK and the Netherlands. Models including between 2 - 4 predictors already provided a discriminative ability comparable to guideline-recommended models: PCE (10 predictors), SCORE2 (8 predictors), and QRISK3 (22 predictors for women, 23 for men).

*Implications of all the available evidence:* We show that even when only a subset of predictor variables is available, our partial models approach can make clinically relevant predictions of the 10-years risk of (major) cardiovascular disease, enabling earlier and more effective treatment decisions. The set of partial models are accessible through a python-based API, allowing for integration in personal or clinical care dashboards.

## INTRODUCTION

Cardiovascular risk prediction models are widely used for treatment initiation and intensification in primary care settings. The UK National Institute for Health and Care Excellence^1^ (NICE) recommends the QRISK3^2^ tool to estimate the 10-year cardiovascular disease (CVD) risk, while the American College of Cardiology/American Heart Association (ACC/AHA) advises using the Pooled Cohort Equations (PCE)^3^ ^4^. In Europe, the European Society of Cardiology suggests the SCORE2^5^ ^6^ model for CVD risk prediction. While these models vary in the number of predictor variables, ranging from 8 in SCORE2 to 22/23 in QRISK3^2^ (see Appendix Table 1), the type of features are relatively comparable and their discriminative ability fairly similar^7^. Their routine application is, however, limited by the need to measure all predictor variables. The absence of even a single predictor - such as high-density lipoprotein cholesterol (HDL-C) in SCORE-2 - renders risk prediction impossible. For example, among individuals between 40 and 69 years without a history of CVD, 71.5% did not have HDL-Cand LDL-C measured at any time across their entire healthcare record; see Appendix Methods and Appendix Table 2. Financial and logistic constraints on testing and screening services, combined with low attendance at general health checks, further reduce opportunities to obtain these missing measurements. As a result, risk prediction remains limited to a small subset of high-risk individuals where complete measurements are actively pursued —most of whom will receive CVD medication—while a much larger population, for whom the full set of predictors is unavailable, is overlooked. Given the relatively comparable performance of many CVD prediction models, which in terms of predictor variables can be viewed as (partial) subset of one another, we sought to develop a set of partial models (considering all combination of 2 to 14 variables) which could be applied in settings where only a subset of predictors was available. The predictive performance of this set of partial models was determined in terms of discrimination and calibration of the 10-years risk of (major) CVD across five cohort studies. Model performance was explored by study as well as across key subgroups: age, sex, socio-economic status, type 2 diabetes, and ethnicity. Feature importance was explored for each study cohort, focusing on the available predictors. Finally, to confirm the model’s relevance in individuals lacking information on canonical cardiovascular risk factors such as blood pressure and blood lipids, we evaluated its performance by masking these variables during external validation.

## METHODS

### Considered predictors for the 10-year risk of major CVD

Predictors were identified by first identifying a set of commonly used ”traditional” CVD predictors based on a prior review and external validation study^7^, as well as their feasibility for measurement in primary care settings. These included smoking status (never, previous, or current smoker), systolic and diastolic blood pressure (SBP/DBP, mmHg), type 2 diabetes (yes/no), the ratio of total cholesterol to HDL-cholesterol, treated hypertension (yes/no), family history of heart disease (yes/no, considering maternal, paternal, and sibling histories)^8^, BMI (kg/m²)^9^. Next this list of traditional predictors was supplemented by a selection of non-traditional predictors which were previously shown to be strongly related to CVD: albumin (g/L)^10^, red blood cell (erythrocyte) distribution width (RDW, %)^11^, and estimated glomerular filtration rate (eGFR, an index of kidney function)^12^ ^13^ . See predictor availability and definitions in the Appendix Table 4, and Appendix Data 1.

### Derivation data

Considering all possible combinations of 12 features (with sex and age always included, to make 14), we derived 4,096 (i.e. 2^12^) separate models. The set of partial models were trained using a random 80% (374,700 individuals) split of the UK Biobank (UKB), selecting individual free of major CVD at the time of enrolment. The models were trained to predict the 10-years risk major CVD, and (externally validated on their ability to predict CVD and major CVD. Here CVD was defined as coronary heart disease (CHD), any stroke, and sudden cardiac death, with major CVD additionally including peripheral artery disease (PAD), atrial fibrillation (AF), or heart failure (HF). The 10-years occurrence of these outcomes was determined through nation-wide linkage of hospital episode statistics (HES) and general practitioner (GP) records, see Appendix Table 3 for outcome definitions. Individuals were followed up from the time of UKB enrolment, until the first occurrence of a major CVD event, non-CVD death, or the end of study, truncating follow-up to 10-years after enrolment, whichever came first. The remaining 20% (93,676 individuals) of the UKB data were used as testing split allowing for unbiased assessments of model performance.

### External validation data

The set of partial models was subsequently externally validated across four fully independent studies from the UK and the Netherlands, specifically we sourced data from the Southall and Brent Revisited^14^ (SABRE) cohort, the Hoorn Study 1 (HS1) ^15^ and Hoorn Study 2 (HS2)^15^ , as well as to the Rotterdam Study^16^ (RS) cohort. Depending on the availability of variables in study cohorts, each individual model of the set of partial models was validated in at least two studies.

The SABRE study is a population-based cohort including White British individuals and first-generation migrants of South Asian or African Caribbean heritage, recruited from West London. The baseline data were collected between 1988 and 1991. In SABRE outcomes were defined based on ICD-9 and ICD-10 codes from death certification, primary care data, HES, and participant reports of physician-diagnosed CHD or stroke^17^ .

The HS^15^ is a population-based cohort study enrolling participants aged 50-75 years between 1989 and 1992 in the Netherlands (HS1) and a second wave enrolling participants aged 40 – 65 years between 2006 and 2007 (HS2). We used questionnaire data to exclude people with prevalent CVD. ICD-9 codes were used to define incident CVD events during a 10-year follow up period.

The RS is a longitudinal population-based cohort, for the current analysis we leveraged information from the RS of people contributing to the 1997-1999 re-examination of the original cohort members. CVD outcome data were collected from the general practitioners and discharge reports from medical specialists, following adjudication by study physicians^18, 19^.

See Appendix Methods, Appendix Table 4, and Appendix Data 1.

### Statistical analysis

The set of 4,096 partial models were derived using a generalized linear model with a binomial distribution and logit link function, and trained to predict the 10-years risk of major CVD. Models were evaluated on discrimination (using Harrell’s C-statistic^20^) and calibration (including calibration-in-the-large (CIL), calibration slope (CS), and calibration plots) considering both CVD and major CVD.

The derivation, testing and external validation cohorts’ variables, were mean centred and standardised to one standard deviation based on the derivation data derived estimates; see Appendix Data 2. To prevent bias during model derivation and validation missing data for variables partially observed (see Table 1) were imputed using Multiple Imputation by Chained Equations (MICE) (single imputation) based on the predictive mean matching algorithm^21^. Variables missing for all study participants were omitted from analysis for the specific cohort i.e. missing at the study level.

Feature importance was assessed using a permutation feature importance algorithm with 10 permutations applied to the test/external validation data, evaluating the change in the c-statistic.

Model performance of the set of partial models was evaluated against the guideline recommended models: PCE^4^ and SCORE2^5^, and QRISK3^2^ using discrimination and calibration metrics; see Application programming interface for models implementation based on the published algorithms. Importantly, we did not attempt to re-estimate the models or model weights, and instead simply applied these models without any model updates – thus mimicking real-world application. The majority of the predictor variables needed for the PCE and SCORE2 application were available in all of the considered cohort studies, with local adjustments described in Appendix Data 1. The QRISK3, which required 20+ variables could only be reliably implemented in the UKB, with five or more QRISK3 variables were missing at the study level in the remaining cohort studies.

### Subgroups analysis

Subgroup performance was explored for age (below or equal and above 65 years), sex, socio-economic status (e.g., Townsend deprivation index or education qualification), type 2 diabetes, and ethnicity, including only those subgroups with at least 10 major CVD. In the UK based studies (UKB and SABRE) the Townsend deprivation index (by tertile) was used as a measure of socio-economic status. Because the Townsend index requires UK postcodes this could not be defined for the Dutch studies (HS1, HS2 and RS), which instead used highest attained educational qualification (primary, secondary, university). Performance by ethnicity was explore using the UKB (European, South Asians, Chinese, Asian or Asian British, African Caribbean, mixed) and SABRE (European, South Asians, African Caribbean). Due to the relatively small number of non-Dutch/European ethnicities enrolled in the HS1, HS2, and RS, similar analyses were impossible in these studies. Differences in discrimination between subgroups were determined using the Wilcoxon test or the Kruskal-Wallis test depending on the number of groups.

### Model performance without considering blood pressure and blood lipids

To determine applicability in individuals without information on the canonical cardiovascular risk factors blood pressure and blood lipids, model performance was additionally explored after masking (i.e. removing) these variables from the testing/external validation data.

### Application programming interface

The application programming interface (API) implementing the set of partial models is available from https://gitlab.com/cvd_in_t2dm/array-of-cvd-prediction-models.

## RESULTS

The training data included 374,700 participants of which 31,590 (8.4%) developed major CVD and 22,664 (6.0%) many CVD, with the testing and external validation data consisting of 105,550 participants of whom 8964 (8.5%) developed major CVD and 6,685 (6.3%) who developed CVD. The available studies reflected populations with distinct patient characteristics, for example the number of women enrolled ranged between 58.9% for the RS and 25% for the SABRE study. The average age ranged between 53.4 for the HS2 and 58.9 for the RS, diabetes ranged between 4.0% in the HS2 to 7.3% in the HS1, treated hypertension ranged between 9.7% in the UKB and 31.5% in SABRE, with family history of heart disease ranging between 21.7% in the HS1 and 41.7% in the UKB, see Table 1.

**Table 1.** Clinical characteristics of the five cohort studies used for validating the set of partial models predicting 10-year risk of major CVD. n.b. Major cardiovascular disease (CVD) represents a composite of coronary heart disease (CHD), ischemic stroke, peripheral artery disease (PAD), atrial fibrillation (AF), and heart failure (HF). SD refers to standard deviation. Abbreviations: body mass index (BMI), diastolic blood pressure (DBP), estimated glomerular filtration rate (eGFR), glycated haemoglobin (HbA1C), high-density lipoprotein cholesterol (HDL cholesterol), systolic blood pressure (SBP). The Kruskal-Wallis test was used to assess statistical differences in continuous variables (e.g., number of individuals, age, BMI, eGFR, albumin, RDW, HDL cholesterol, total cholesterol, HbA1c, SBP, and DBP) across studies, while the Chi-squared test was applied for categorical variables (e.g., smoking status, T2DM, family history of heart disease, treated hypertension, and Major CVD outcomes).

|  | UK Biobank<br>(training dataset) |  | UK Biobank<br>(testing dataset) |  | SABRE<br>(independent validation) |  | Rotterdam Study<br>(independent validation) |  | The Hoorn Study 1<br>(independent validation) |  | The Hoorn Study 2<br>(independent validation) |  | P-value<br>(difference between testing and external validation study cohorts) |
| --- | --- | --- | --- | --- | --- | --- | --- | --- | --- | --- | --- | --- | --- |
| Clinical characteristic | Mean (SD) or N (%) | Missing data (%) | Mean (SD) or N (%) | Missing data (%) | Mean (SD) or N (%) | Missing data (%) | Mean (SD) or N (%) | Missing data (%) | Mean (SD) or N (%) | Missing data (%) | Mean (SD) or N (%) | Missing data (%) |  |
| Total no. of individuals | 374,700 |  | 93,676 |  | 3,861 |  | 3,282 |  | 2,248 |  | 2,483 |  |  |
| Women (%) | 209,930 (56.0) | 0.0 | 52,253 (55.8) | 0.0 | 967 (25.0) | 0.0 | 1,932 (58.9) | 0.0 | 1,250 (55.6) | 0.0 | 1,326 (53.4) | 0.0 | $1.00 \times 10^{-100}$ |
| Age (years) | 56.2 (8.1) | 0.0 | 56.3 (8.1) | 0.0 | 52.0 (6.8) | 0.0 | 56.5 (6.6) | 0.0 | 61.5 (7.4) | 0.0 | 53.3 (6.7) | 0.0 | $3.59 \times 10^{-10}$ |
| Smoking Status | | 0.6 | | 0.6 | | 0.3 | | 0.3 | | 0.8 | | 0.6 | $1.00 \times 10^{-100}$ |
| Never smoker | 208,200 (55.6) |  | 51,812 (55.3) |  | 2,104 (54.5) |  | 1,018 (31.0) |  | 793 (35.3) |  | 971 (39.1) |  |  |
| Previous smoker | 125,529 (33.5) |  | 31,428 (33.5) |  | 853 (22.1) |  | 1,386 (42.2) |  | 737 (32.8) |  | 986 (39.7) |  |  |
| Current smoker | 38,881 (10.4) |  | 9,905 (10.6) |  | 894 (23.2) |  | 869 (26.5) |  | 699 (31.1) |  | 512 (20.6) |  |  |
| BMI (kg/m2) | 27.3 (4.7) | 0.5 | 27.3 (4.7) | 0.5 | 26.4 (4.0) | 0.1 | 27.7 (4.6) | 0.8 | 26.5 (3.6) | 0.2 | 26.2 (4.0) | 1.4 | $3.59 \times 10^{-10}$ |
| eGFR | 95.0 (12.9) | 6.4 | 95.1 (12.8) | 6.4 | 97.3 (16.7) | 33.6 | 90.2 (13.1) | 3.8 | 73.7 (12.6) | 0.2 | N/A | N/A | $7.58 \times 10^{-8}$ |
| Albumin (g/L) | 45.2 (2.6) | 14.4 | 45.2 (2.6) | 14.4 | N/A | N/A | N/A | | 38.5 (2.8) | 0.2 | N/A | N/A | $8.09 \times 10^{-3}$ |
| Red blood cell distribution width (RDW) (%) | 13.5 (1.0) | 4.8 | 13.5 (1.0) | 4.9 | N/A | N/A | 12.3 (0.9) | 0.9 | N/A | N/A | N/A | N/A | $8.05 \times 10^{-3}$ |
| HDL cholesterol (mmol/L) | 1.5 (0.5) | 12.7 | 1.5 (0.4) | 12.7 | 1.4 (0.4) | 16.2 | 1.4 (0.4) | 1.2 | 1.3 (0.4) | 0.3 | 1.5 (0.4) | 0.2 | $3.59 \times 10^{-10}$ |
| Total cholesterol (mmol/L) | 5.8 (1.1) | 6.3 | 5.8 (1.1) | 6.3 | 6.0 (1.1) | 0.7 | 5.6 (1.0) | 1.2 | 6.6 (1.2) | 0.2 | 5.5 (1.0) | 0.2 | $3.59 \times 10^{-10}$ |
| HbA1C (mmol/mol) | 35.9 (6.3) | 7.1 | 35.9 (6.4) | 7.0 | 40.6 (11.2) | 36.1 | N/A | | 36.4 (9.3) | 0.3 | 36.0 (5.3) | 0.2 | $7.66 \times 10^{-8}$ |
| SBP (mm Hg) | 139.7 (19.7) | 0.2 | 139.7 (19.6) | 0.2 | 124.3 (17.3) | 0.1 | 132.5 (19.0) | 0.5 | 135.2 (20.2) | 0.1 | 133.1 (17.7) | 1.5 | $3.59 \times 10^{-10}$ |
| DBP (mm Hg) | 82.3 (10.6) | 5.9 | 82.3 (10.6) | 5.8 | 78.8 (11.0) | 0.1 | 82.8 (11.0) | 0.5 | 82.1 (10.4) | 0.4 | 76.5 (10.7) | 1.5 | $3.59 \times 10^{-10}$ |
| Type 2 diabetes (%) | 25,005 (6.7) | | 6,337 (6.8) | | 250 (6.5) | 0.0 | 179 (5.5) | | 163 (7.3) | | 99 (4.0) | | $5.89 \times 10^{-8}$ |
| Family history of heart disease (%) | 156,245 (41.7) | | 39,092 (41.7) | | N/A | N/A | 718 (21.9) | | 488 (21.7) | | 589 (23.7) | | $1.00 \times 10^{-100}$ |
| Treated hypertension (%) | 36,678 (9.8) | | 9,128 (9.7) | | 425 (22.1) | 0.0 | 1,033 (31.5) | | 357 (15.9) | | 591 (23.7) | | $1.00 \times 10^{-100}$ |
| **Outcomes** |  |  |  |  |  |  |  |  |  |  |  |  |  |
| Major CVD (%) | 31,590 (8.4) | | 7894 (8.4) | | 388 (10.1%) | | 158 (4.8%) | | 359 (16.0%) | | 165 (6.7%) | | $2.92 \times 10^{-51}$ |

While all the variables used in the set of partial models were available in the UKB data (used for training and testing), following our intended application the availability of predictors differed across the external validation studies: the SABRE study did not record information on albumin, RDW, and family history of heart disease, the RS did not include information on albumin and HbA1c, the HS1 lacked data on RDW, and the HS2 did not have information on eGFR, albumin, RDW; see Appendix Table 4.

### Model performance in the training, testing, and external validation data

Model performance was near constant irrespective of the considered outcome: major CVD or CVD, hence in the following we focussed on major CVD; See Appendix Tables 5-6 and Appendix Figures 1–2. Training discrimination ranged between a c-statistic of 0.70 (95% CI 0.70; 0.70) for 2 predictors (age and sex only) to 0.73 (95% CI 0.73; 0.73) for 14 variables; see Appendix Data 3. The UKB testing performance showed identical results with a c-statistic between 0.70 (95% CI 0.70, 0.70) for 2 features (age and sex only) up to 0.73 (95% CI 0.73, 0.73) for all available predictors (n = 14). In the UKB testing data the partial models were reasonably well-calibrated, with UKB training CS ranging from 1.09 (95% CI 1.05; 1.13) for 4 variables to 1.14 (95% CI 1.10; 1.17) for 13 predictors, and the testing CIL ranging between 0.12 (95% CI 0.10; 0.15) and 0.14 (95% CI 0.11; 0.16); see Figures 1-2, Appendix Data 3, Appendix Figures 3–4, and Appendix Tables 7-8.

**Figure 1.**
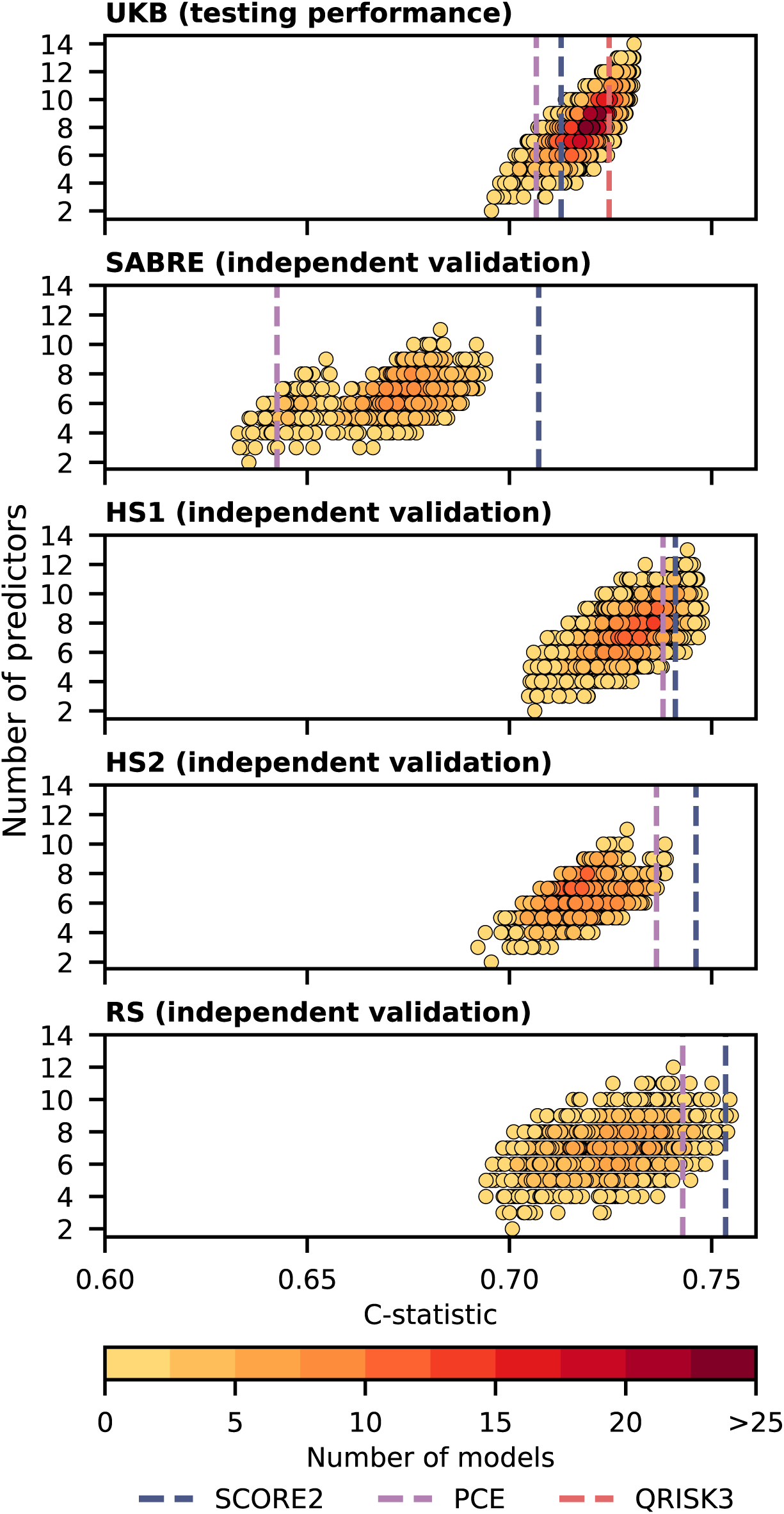
Discriminative performance of the set of partial models predicting 10-year risk of major CVD across five cohort studies. n.b. The discrimination (C-statistic, x-axis) is plotted against the number of predictors included in the major CVD risk prediction models. The set of partial models is compared to established CVD risk prediction models, including SCORE2, PCE, and QRISK3 (represented by vertical dotted lines). See discrimination metrics in the Appendix Data 3.

**Figure 2.**
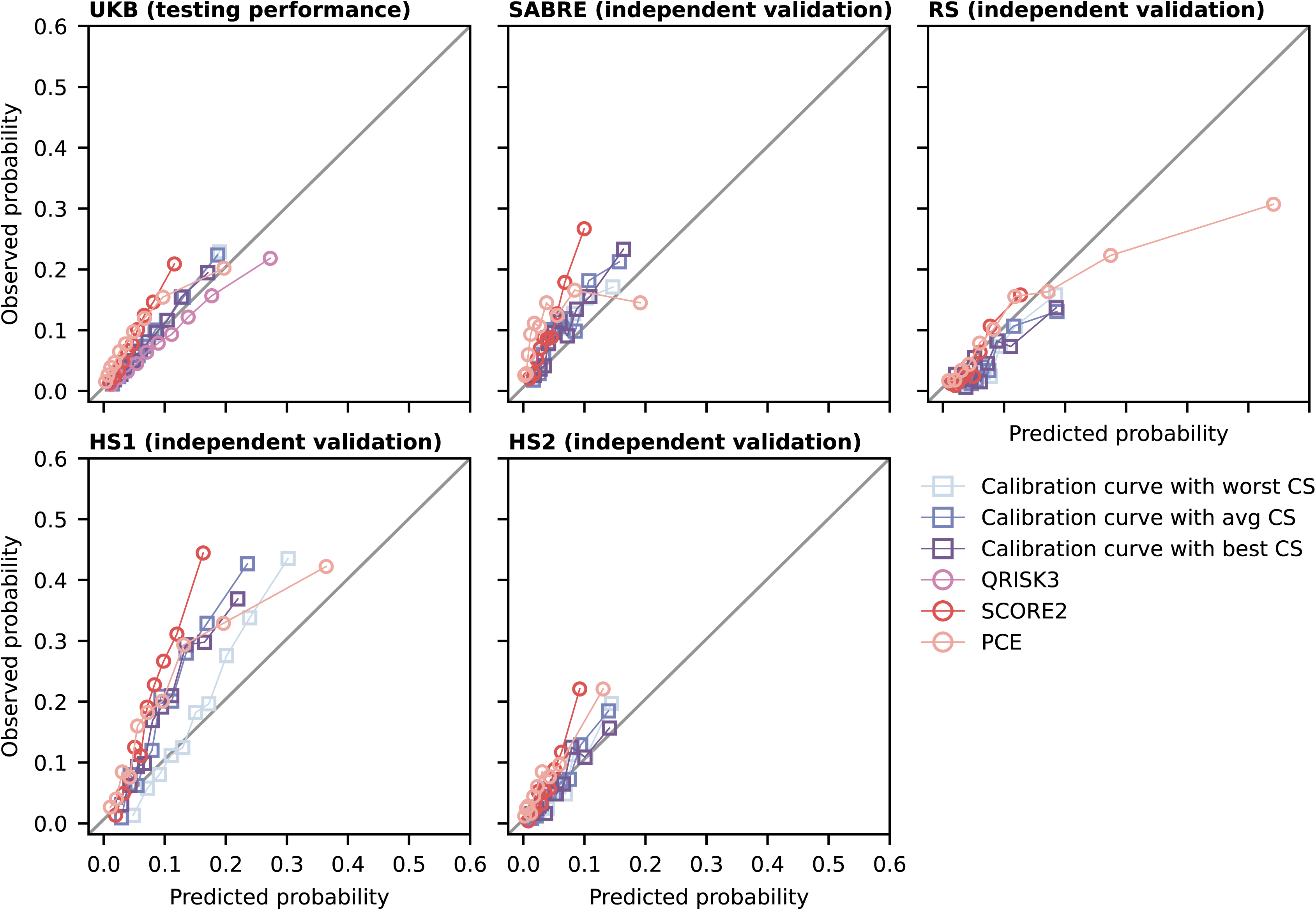
Calibration plots for selected models from the set of partial models predicting the 10-year risk of major CVD across five cohort studies. n.b. Calibration is presented as the minimal, average, and maximal values of calibration slopes, are shown alongside the calibration of established CVD risk prediction models like SCORE2, PCE, and QRISK3. The models’ calibration was assessed in five study cohorts: the UK Biobank (UKB), SABRE, Hoorn Study 1 (HS1), Hoorn Study 2 (HS2), and the Rotterdam Study (RS). The observed 10-year risk (y-axis) is plotted against the mean predicted 10-year risk (x-axis) within groups defined by quantiles of predicted risk. The diagonal line indicates perfect calibration. See calibration metrics in the Appendix Data 4.

**Figure 3.**
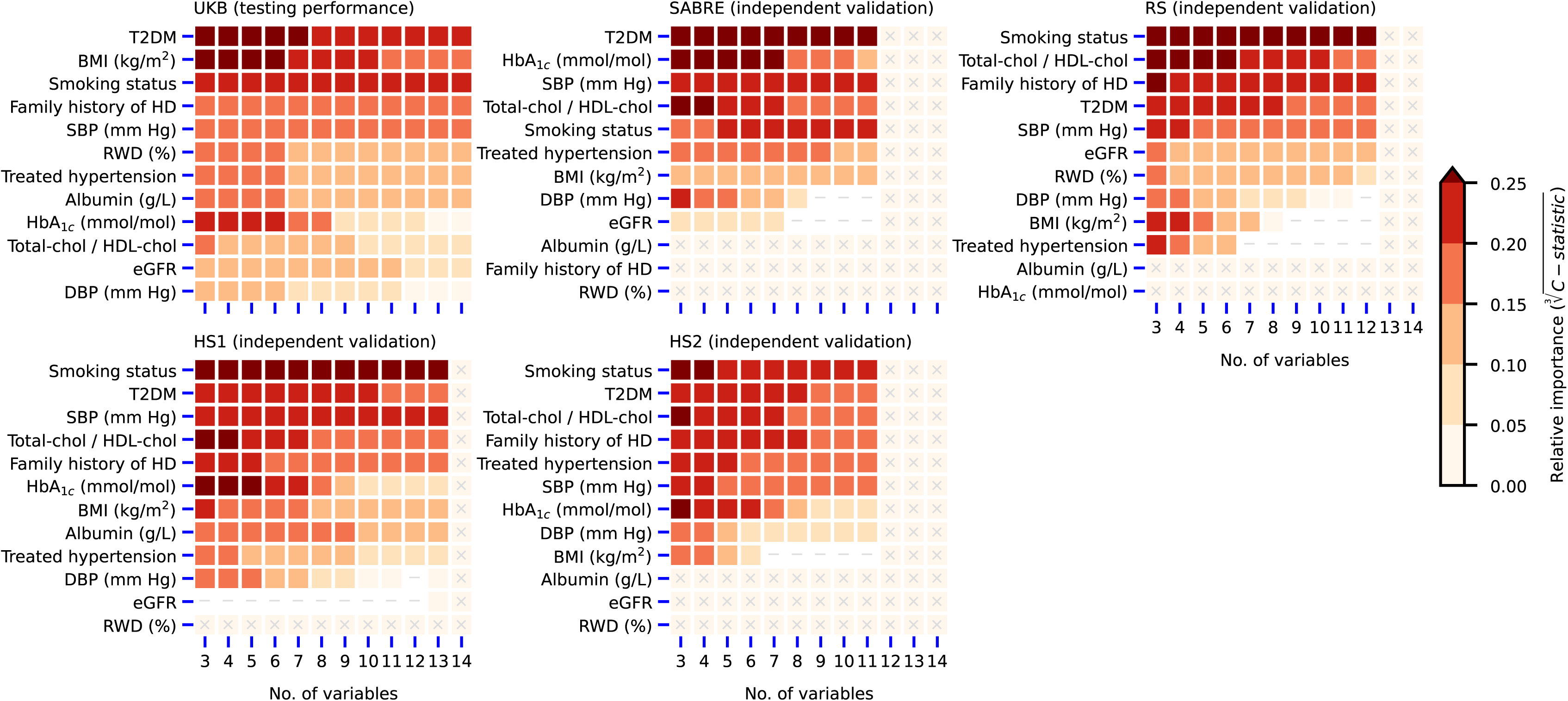
The median feature importance for the set of partial models predicting 10-year risk of major CVD evaluated across five cohort studies. n.b. The median feature importance for the set of partial models predicting 10-year risk of major CVD evaluated across five study cohorts. The permutation feature importance assesses the c-statistic change in the study cohort; iteratively the values of each predictor were randomly assigned to an individual after which the c-statistic was re-estimated with these permuted data and the difference in performance used as an estimate of predictor contribution to the model’s predictive potential. The x-axis shows the features considered in the prediction models, while the y-axis represents the number of variables included in the set of partial models predicting 10-year risk of major CVD. The colour indicates the median importance of predictor features across the set of partial models using a specific number of predictors. Cells marked with a cross indicate variables not considered by the partial models due to their unavailability in the study cohort, and cells marked with a minus sign indicate variables with a negative median feature importance. See feature importance in the Appendix Data 5.

**Figure 4.**
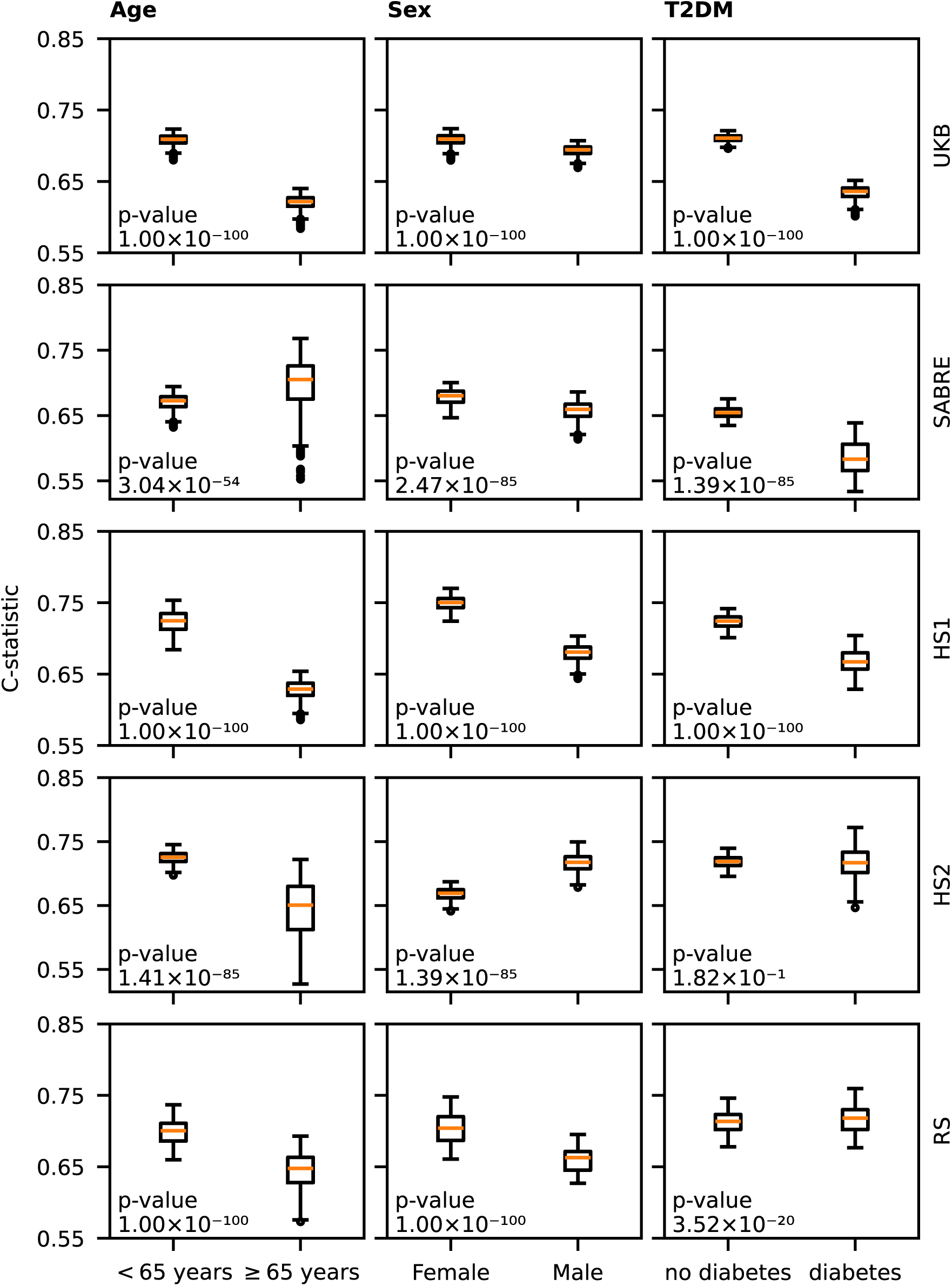
Discriminative performance of the set of partial models predicting 10-year risk of major CVD per study participant subgroups. n.b. Discriminative performance of the set of partial models predicting 10-year risk of major CVD per UK Biobank (UKB), Southall and Brent Revisited (SABRE), Hoorn Study 1 (HS1), Hoorn Study 2 (HS2) and Rotterdam Study (RS) participant subgroups. The p-value is calculated using the Wilcoxon test for subgroups with two categories and the Kruskal-Wallis test for subgroups with more than two categories. See participant subgroup sizes in Appendix Table 9, the proportion of major CVD events in Appendix Table 10, and subgroup definitions in the Appendix Data 1.

The external validation results showed somewhat improved discrimination in the RS (max c-statistic 0.75 (95% CI 0.75; 0.76)), HS1 (max c-statistic 0.75 (95%CI 0.75; 0.75)), and HS2 (max c-statistic 0.74 (95% CI 0.74; 0.74)), and attenuated performance in the more ethnically diverse SABRE (max c-statistic 0.69 (95% CI 0.66; 0.72)). The partial models were reasonably well calibrated in the RS and HS2, while moderately underestimating the risk of major CVD in the SABRE and OHS; Figure 2, Appendix Tables 7-8, Appendix Data 3-4.

### Feature importance

Exploring feature importance across five cohorts confirmed that age and sex are crucial variables for major CVD risk prediction, consistently ranking among the top two features in most cohorts; see Appendix Figure 5. Due to correlation of age and sex with the 12 features that made up the partial models, the importance of age and sex attenuated as the number of considered features increased.

As anticipated feature importance changed with increasing number of model parameters, as well as across the different studies. For example, in the UKB testing set (Figure 3) diabetes, HbA1c, and BMI strongly contributed to the models discriminative ability when were between 2 and 6/7 features, but their contribution decreases in models using additional features. In SABRE, type 2 diabetes and HbA1c were ranked higher than sex in 340 out of 512 models, where type 2 diabetes remained important irrespective of the number of available features. This may be partially attributed to the skewed sex distribution in the SABRE cohort, with a male-to-female ratio of 3:1. In the HS1 smoking status was a consistently strong predictor, with T2DM and HbA1c losing importance in the models using additional features. In the HS2 smoking status, the cholesterol/HDL-C ratio, and HbA1c were particularly important features in partial models with 3 features but decreased in importance in more complicated models. Finally, in the RS smoking status, the cholesterol/HDL-C ratio, and family history of heart disease were important features to predict major CVD; please see Figure 3 and Appendix Data 5.

### Comparison against guidelines recommended CVD prediction models

In the testing sample of the UKB, the partial models showed a similar discriminative ability as the guideline recommended models, for example the QRISK reached a c-statistic of 0.72 (95% CI 0.72, 0.73).

Across the four external validation cohorts the set of partial models generally outperformed the PCE, reaching discriminative abilities equal or close to that of the SCORE2. Here the QRISK3 could not be evaluated due to a lack of predictor variables in the cohort used for external validation. While calibration was highly accurate in the UKB testing data, agreement between observed and predicted risk slightly decreased in external validation settings, which generally equalled that of the guideline recommended models.); see Figures 1-2, Appendix Figures 3–4 and Appendix Table 5.

### Subgroup analysis

Subgroup analyses indicated that the set of partial models was slightly more accurate in people aged 65 or less (p-values for the difference <0.05) in all cohort studies aside from SABRE. Aside from the HS2 study, we found that the set of partial models performed better in women than in men (p-values for the difference < 0.05) and median difference across studies between 0.02 and 0.07; see Figure 4. Additionally, we observed attenuated performance in people with T2DM in most studies (median difference between 0.005 and 0.075), expect for the HS2. We did not observe a meaningful difference between participants based on deprivation status, education, or ethnicity; see Appendix Tables 9–10, and Appendix Figures 6 – 10.

The guideline recommended models showed a similar subgroup specific performance, with small-to-moderately attenuated performance in men, people aged over 65 years, and people with T2DM. We did observe some model specific differences, with for example PCE showing decreased performance between ethnicities in the two UK-based studies (SABRE and UKB), while there was no strong evidence for differential performance of QRISK3 (only evaluated in the UKB) and SCORE2 in these settings; see Appendix Data 6.

### Performance when masking blood pressure and blood lipids

We additionally evaluated the discriminative ability of the set of partial model when blood pressure and blood lipids were masked. This generally results in similar performance (Q1/Q3 UKB 0.71/0.72, SABRE 0.66/0.67, HS1 0.72/0.73, HS2 0.71/0.72, RS 0.71/0.73) compared to performance without masking (Q1/Q3 UKB 0.71/0.73, SABRE 0.67/0.68, HS1 0.73/0.74, HS2 0.71/0.73, RS 0.72/0.74); see Appendix Table 11.

## Discussion

Routine CVD risk prediction is limited to people with complete information on all model variables, which may limit risk prediction to people with a clinically suspected higher risk, rather than the entire population. To increase applicability of risk prediction models for the entire population, we have developed a straightforward partial models solution, training 4,096 models using different combinations of 14 variables. Importantly across the five considered cohort studies the set of partial models generally performed comparably to the guidelines recommended models PCE (9 variables), SCORE (8 variables) and QRISK (22 variables). To ensure wider applicability, the set of partial models were developed to include a combination of traditional risk factors (e.g. blood lipids, blood pressure, smoking) and non-traditional risk factors (e.g. HbA1c, albumin, RWD). In line with this we show that the subset models without blood lipids and blood pressure provide a similar discriminative performance (512 without this information compared to 3584 including these variables) further supporting application in people without measurements of these canonical CVD risk factors.

While predict performance of the set of partial models improved when increasing the number features, relatively small models using 2-4 predictors nevertheless reached a c-statistic of around 0.70 and did not markedly under-perform compared to the considered guideline recommended models. Irrespective of the number of features, the set of partial models showed equally reasonable calibration as compared to the considered guidelines recommend models. Subgroup analyses revealed modest differences in model performance by age, sex and diabetes status for both the set of partial and guideline-recommended models, where performance improved in lower risk groups (e.g., in people aged 65 years or younger, women, and people without diabetes), which aligns with prior studies^2^ ^7^.

The set of features considered by the partial models was pre-selected to combine traditional and non-traditional risk factors, allowing for a broad range of application, even in settings were traditional risk factors (e.g. cholesterol measurements) were not-necessarily available. To enhance applicability to individuals without a suspected high risk of cardiovascular disease, we specifically evaluated the performance of models that exclude information on blood pressure and blood lipids. The finding that this reduced set of models performs equally well suggests that accurate prediction of major CVD risk is possible without additional laboratory measurements - potentially lowering the threshold for uptake. Applying the set of partial models across five cohort studies furthermore indicated that a different set of features was relevant in each study, revealing the flexibility of our approach.

The set of partial models was trained to predict major CVD consisting of CVD, AF, HF, and PAD. Using the testing and external validation data we show that performance of the set of partial models is equal when predicting a more traditional definition of CVD, exclusively considering the composite of CHD, stroke, and sudden cardiac death.

The key strength of this study includes the development of a comprehensive set of 4,096 partial models for predicting the 10-year risk of major CVD, which allows for risk estimation using only two predictors (age and sex). The models’ performance improves as additional features are added, approaching performance of guideline-recommended models. This highlights that even with partial data, estimated major CVD risk can provide valuable clinical guidance. The set of major CVD partial models was thoroughly validated across five cohorts, each comprising distinct patient populations, enabling the identification of both common and unique features important in each study.

We applied imputation to ensure each model was evaluated using the same participants, and as such that differences in performance was due to the considered features rather than the participants without missing data. By doing this show that in real-world application, where such imputation is unavailable and instead the model with maximum number of features is used, these models perform sufficiently to inform shared medical decisions. While our approach of using partial models provides reasonably accurate predictions, should there be sufficient information to use one of the guideline recommended models this is likely to provide similar or better predictions. However, as discussed in the available EHR less than 30% of the participants aged 40-69 had a an ever measurement of HDL-C and LDL-C, without which none of the considered guideline recommended models based be employed. Our aim is not to replace these models, but to offer a reasonably accurate alternative for situations where acquiring the additional measurements required by guideline-recommended models is impractical due to patient burden, cost, or time constraints. To facilitate implementation of our set of partial models we have developed an API which can be included in clinical software or online patient dashboards, and which automatically returns the predicted risk within milliseconds for appropriate partial model which uses all of the supplied features.

Among the limitations of this study is the restricted number of predictors considered for developing the set of major CVD partial models, primarily due to computational constraints at time of derivation. We do note that including additional features only incrementally improved discriminative performance, and as such more complex models may not always offer meaningful improvements. Additionally, due to the unavailability of certain variables in specific cohorts, it was not feasible to validate every model in all five study cohorts; however, each model was validated in at least two cohorts. Similarly, while we had access to detailed individual participant records allowing for necessary data harmonisations, the UKB based studies exclusively enrolled sufficient non-European participants to explore potential difference by ethnicity. While this analysis did not find support for meaningful difference between the different ethnic groups, we were unable to perform similar analyses in the Dutch cohorts.

In summary, the set of 4,096 validated partial models for predicting the 10-year risk of major CVD offers the potential for accurate risk prediction and discrimination in settings where due to missing information application of a guideline recommended CVD prediction model is impracticable. These set of partial models has been thoroughly validated across five independent study cohorts. The open software API facilitates straightforward application and validation of the partial models in healthcare settings.

## Contributors

AFS, FWA, NC contributed to the idea and design of this study. KD prepared the datasets for analysis, derived multiple partial models, and performed external validation. SVE, JWJB, PPH, DB, and ML provided data access and local coding support for external validation. KD drafted the initial version of the manuscript. SVE, JWJB, PPH, ML, DB, MK, FWA, NC, and AFS provided critical feedback on the analysis, its interpretations, and commented on the drafted manuscript. KD is responsible for the integrity of the work as a whole.

## Data sharing

The UK Biobank data are available pending an approved application. Data from the remaining cohort are available, pending ethical approvals, from the local principle investigators.

Analyses were carried out in Python v3.6 using *scikit-learn*^22^, *statsmodels*^23^, *pandas*^24^, and *numpy*^25^, plots were generated using *matplotlib*^26^, and *seaborn*^27^, imputation was performed in R v4.1 using *mice*^21^. See https://gitlab.com/cvd_in_t2dm/array-of-cvd-prediction-models for the API and codebase underpinning this work.

## Acknowledgements

This work is partially supported by MyDigiTwin with project number 628.011.213 of the research programme “COMMIT2DATA – Big Data & Health” which is partly financed by the Dutch Research Council (NWO). AFS is supported by BHF grants PG/18/5033837, PG/22/10989, the UCL BHF Research Accelerator AA/18/6/34223. AFS and SVE received additional support from the National Institute for Health Research University College London Hospitals Biomedical Research Centre. This work was funded by UK Research and Innovation (UKRI) under the UK government’s Horizon Europe funding guarantee EP/Z000211/1, by the UKRI/NIHR Multimorbidity fund Mechanism and Therapeutics Research Collaborative MR/V033867/1, and by the Rosetrees Trust. This publication is part of the project “Computational medicine for cardiac disease” with file number 2023.022 of the research programme “Computing Time on National Computer Facilities” which is (partly) financed by the Dutch Research Council (NWO). The authors acknowledge the use of the UCL Myriad High Performance Computing Facility (Myriad@UCL), and associated support services, in the completion of this work.

This research has been conducted using the UK Biobank Resource under application numbers 12113, 24711 and 44972. We are grateful to the UK Biobank participants. UK Biobank was established by the Wellcome Trust medical charity, Medical Research Council, Department of Health, Scottish Government, and the Northwest Regional Development Agency. It has also had funding from the Welsh Assembly Government and the British Heart Foundation. SABRE was funded by the Wellcome Trust, British Heart Foundation, Diabetes UK and the Medical Research Council.

The Rotterdam Study is funded by the Erasmus MC – University Medical Center Rotterdam and the Erasmus University Rotterdam, the Netherlands Organization for the Health Research and Development (ZonMw), the Research Institute for Diseases in the Elderly (RIDE), the Ministry of Education, Culture and Science, the Ministry for Health, Welfare and Sports, the European Commission (DG XII), and the Municipality of Rotterdam. The Rotterdam Study has been approved by the Medical Ethics Committee of the Erasmus MC (registration number MEC 02.1015) and by the Dutch Ministry of Health, Welfare and Sport (Population Screening Act WBO, license number 1071272-159521-PG). The Rotterdam Study has been entered into the Netherlands National Trial Register (NTR; www.trialregister.nl) and into the WHO International Clinical Trials Registry Platform (ICTRP; www.who.int/ictrp/network/primary/en/) under shared catalogue number NTR6831. All participants provided written informed consent to participate in the study and to have their information obtained from treating physicians.

Funding for the Hoorn Study was provided by the VU University Medical Center of Amsterdam, the Dutch Research Council (NWO), the Netherlands Organization for Health Research and Development (ZonMW), the Dutch Diabetes Research Foundation and the Dutch Heart Foundation. Funding for the New Hoorn Study was provided by: the VU University Medical Center of Amsterdam, Novartis Pharma B.V, the European Union and the Innovative Medicine Initiative.

The authors are grateful to the study participants, the staff from the Rotterdam Study and the participating general practitioners and pharmacists. We appreciate the corporation of the participants and research assistants who have been involved in the Hoorn Study and New Hoorn Study.

## Prior postings and presentations

This study and its results have not been published previously.

