## Appendix for "Accurate machine learning-based CVD risk prediction in primary care may reduce the need for routine health care checks"

6 Discriminative performance of the set of partial models predicting 10-year risk of major CVD per UK Biobank participant subgroup. . 26
7 Discriminative performance of the set of partial models predicting 10-year risk of major CVD per SABRE participant subgroup. . . . . 26
8 Discriminative performance of the set of partial models predicting 10-year risk of major CVD per Hoorn Study 1 participant subgroup. 27
9 Discriminative performance of the set of partial models predicting 10-year risk of major CVD per Hoorn Study 2 participant subgroup. 27
10 Discriminative performance of the set of partial models predicting 10-year risk of major CVD per Rotterdam Study participant subgroup. 28

### Appendix

#### 1.1 Methods

##### 1.1.1 Outcome definitions

CVD was defined as the occurrence of fatal or non-fatal myocardial infarction (MI), sudden cardiac death, ischemic heart disease, fatal or non-fatal stroke or PAD after the start of follow-up. The definition of CVD also includes heart failure (HF) and/or atrial fibrillation (AF), as well as the individual CVD components: CHD, stroke, AF, and HF; see Appendix Table 3.

##### 1.1.2 Derivation data

Data was sourced from the UKB, a cohort of  500,000 men and women aged 40-69 years between 2006 and 2010 enrolled from primary care registers across the UK. Health-related outcomes were defined using the Clinical Practice Research Datalink (CPRD), including primary care data, Hospital Episode Statistics (HES), and mortality data.

##### 1.1.3 Electronic Healthcare Records

To evaluate the extent of missing data in electronic healthcare records, the availability of total cholesterol and high-density lipoprotein (HDL) cholesterol was assessed in the UKB cohort, excluding individuals with CVD at baseline. Measurements of total cholesterol and HDL cholesterol extracted from primary care data were coded using Read Version 2 (Read2) and Read Version 3 (Clinical Terms 3 or CTV3). See Appendix Table 2 for variable definitions.

#### 1.2 Tables

**Table 1:** Predictors used in PCE, QRISK3, and SCORE2 risk models

| Predictor | PCE | QRISK3 | SCORE2 |
|---|---|---|---|
| Age | ✓ | ✓ | ✓ |
| Sex | ✓ | ✓ | ✓ |
| Smoking Status | ✓ | ✓ | ✓ |
| Systolic Blood Pressure (SBP) | ✓ | ✓ | ✓ |
| Diastolic Blood Pressure (DBP) | ✓ | ✓ | |
| Total Cholesterol | ✓ | ✓ | ✓ |
| HDL Cholesterol | ✓ | ✓ | ✓ |
| Type 2 Diabetes | ✓ | ✓ | ✓ |
| Body Mass Index (BMI) | | ✓ | |
| Ethnicity | ✓ | ✓ | |
| Family History of CVD | | ✓ | |
| Chronic Kidney Disease (CKD) | | ✓ | |
| Rheumatoid Arthritis (RA) | | ✓ | |
| Atrial Fibrillation | | ✓ | |
| Townsend Social Deprivation Index | | ✓ | |
| Treated hypertension | ✓ | ✓ | |
| Type 1 Diabetes | | ✓ | |
| Migraine | | ✓ | |
| Corticosteroid use | | ✓ | |
| Systemic lupus | | ✓ | |
| Atypical antipsychotic medication | | ✓ | |
| Severe mental illness | | ✓ | |
| Erectile dysfunction (in men) | | ✓ | |
| Region-Specific Risk Factors | | | ✓ |
| Total number of predictors | 10 | women: 22<br>men: 23 | 8 |

**Table 2:** Definitions of primary care data variables based on Read Code Version 2 and Read Code Version 3.

| Measurement name | Read Codes version 2 and 3 |
|---|---|
| Total Cholesterol | 44OE. Plasma total cholesterol level<br>44P1. Serum cholesterol normal<br>44P2. Serum cholesterol borderline<br>44P3. Serum cholesterol raised<br>44P4. Serum cholesterol very high<br>44P.. Serum cholesterol<br>44P9. Serum cholesterol studies<br>44PH. Total cholesterol measurement<br>44PJ. Serum total cholesterol level<br>44PZ. Serum cholesterol NOS<br>662a. Cholesterol |
| HDL Cholesterol | 44d2. Plasma random HDL cholesterol level<br>44dA. Plasma HDL cholesterol level<br>44P5. Serum HDL cholesterol level<br>44PC. Serum random HDL cholesterol level |

**Table 3:** Outcome definitions based on the Health Data Research (HDR) UK Phenotype Library.

| Type of cardiovascular disease | Link to the HDR UK Phenotype Library |
|---|---|
| Fatal and non-fatal MI | https://phenotypes.healthdatagateway.org/phenotypes/PH215/version/430/detail/<br>https://phenotypes.healthdatagateway.org/phenotypes/PH1027/version/2263/detail/ |
| Fatal or non-fatal Stroke | https://phenotypes.healthdatagateway.org/phenotypes/PH55/version/110/detail/<br>https://phenotypes.healthdatagateway.org/phenotypes/PH56/version/112/detail/<br>https://phenotypes.healthdatagateway.org/phenotypes/PH85/version/170/detail/<br>https://phenotypes.healthdatagateway.org/phenotypes/PH86/version/172/detail/ |
| Fatal peripheral vascular disease | https://phenotypes.healthdatagateway.org/phenotypes/PH236/version/472/detail/ |
| Sudden Cardiac Death | https://phenotypes.healthdatagateway.org/phenotypes/PH995/version/2173/detail/ |
| HF | https://phenotypes.healthdatagateway.org/phenotypes/PH1028/version/2265/detail/ |
| AF | https://phenotypes.healthdatagateway.org/phenotypes/PH36/version/72/detail/ |
| Ischemic stroke | https://phenotypes.healthdatagateway.org/phenotypes/PH56/version/112/detail/,<br>https://phenotypes.healthdatagateway.org/phenotypes/PH85/version/170/detail/ |
| Haemorrhagic stroke | https://phenotypes.healthdatagateway.org/phenotypes/PH55/version/110/detail/,<br>https://phenotypes.healthdatagateway.org/phenotypes/PH86/version/172/detail/ |

**Table 4:** Availability of predictor variables across five study cohorts and the number of partial models considered for predicting the 10-year risk of cardiovascular disease.

| Clinical characteristic | UK Biobank | SABRE | Rotterdam Study | Hoorn Study 1 | Hoorn Study 2 |
|---|---|---|---|---|---|
| Sex (Women/Men) | ✓ | ✓ | ✓ | ✓ | ✓ |
| Age (years) | ✓ | ✓ | ✓ | ✓ | ✓ |
| Smoking Status (Never/Previous/Current) | ✓ | ✓ | ✓ | ✓ | ✓ |
| BMI (kg/m2) | ✓ | ✓ | ✓ | ✓ | ✓ |
| eGFR | ✓ | ✓ | ✓ | ✓ | |
| Albumin (g/L) | ✓ | | | ✓ | |
| Red blood cell distribution width (RDW) (%) | ✓ | | ✓ | | |
| HDL cholesterol (mmol/L) | ✓ | ✓ | ✓ | ✓ | ✓ |
| Total cholesterol (mmol/L) | ✓ | ✓ | ✓ | ✓ | ✓ |
| HbA1C (mmol/mol) | ✓ | ✓ | | ✓ | ✓ |
| SBP (mm Hg) | ✓ | ✓ | ✓ | ✓ | ✓ |
| DBP (mm Hg) | ✓ | ✓ | ✓ | ✓ | ✓ |
| Type 2 diabetes (%) | ✓ | ✓ | ✓ | ✓ | ✓ |
| Family history of heart disease (%) | ✓ | | ✓ | ✓ | ✓ |
| Treated hypertension (%) | ✓ | ✓ | ✓ | ✓ | ✓ |
| No. of considered CVD partial models | 4096 | 512 | 1024 | 2048 | 512 |

n.b. Abbreviations: major cardiovascular disease (CVD) represents a composite of coronary heart disease (CHD), ischemic stroke, peripheral artery disease (PAD), atrial fibrillation (AF), and heart failure (HF), body mass index (BMI), estimated glomerular filtration rate (eGFR), red blood cell distribution width (RDW),high-density lipoprotein cholesterol (HDL cholesterol), glycated haemoglobin (HbA1C), systolic blood pressure (SBP), diastolic blood pressure (DBP).

**Table 5:** External validation of three guideline recommended prediction models for the 10-year risk of major CVD.

| Model | No. predictors | Study | Total sample size | No. of major CVD events (%) | C-statistic | CS | CIL |
|---|---|---|---|---|---|---|---|
| PCE | 9 | UK Biobank | 93676 | 7894 (8.4) | 0.71 (0.70; 0.71) | 0.60 (0.58; 0.62) | 0.59 (0.57; 0.62) |
| PCE | 9 | SABRE | 3861 | 388 (10.1%) | 0.64 (0.61; 0.67) | 0.36 (0.29; 0.44) | 1.02 (0.90; 1.13) |
| PCE | 9 | Hoorn Study 1 | 2248 | 359 (16.0%) | 0.74 (0.74; 0.74) | 0.78 (0.67; 0.89) | 0.79 (0.67; 0.90) |
| PCE | 9 | Hoorn Study 2 | 2483 | 165 (6.7%) | 0.74 (0.73; 0.74) | 0.71 (0.56; 0.86) | 0.80 (0.63; 0.97) |
| PCE | 9 | Rotterdam Study | 3282 | 158 (4.8%) | 0.74 (0.74; 0.74) | 0.62 (0.57; 0.67) | -0.25 (-0.32; -0.17) |
| QRISK3 | 22 | UK Biobank | 93676 | 7894 (8.4) | 0.72 (0.72; 0.73) | 0.94 (0.91; 0.97) | -0.19 (-0.21; -0.17) |
| SCORE2 | 8 | UK Biobank | 93676 | 7894 (8.4) | 0.71 (0.71; 0.72) | 1.18 (1.14; 1.22) | 0.60 (0.58; 0.62) |
| SCORE2 | 8 | SABRE | 3861 | 388 (10.1%) | 0.71 (0.68; 0.74) | 1.24 (1.06; 1.43) | 0.97 (0.86; 1.07) |
| SCORE2 | 8 | Hoorn Study 1 | 2248 | 359 (16.0%) | 0.74 (0.74; 0.74) | 1.55 (1.33; 1.76) | 1.06 (0.95; 1.17) |
| SCORE2 | 8 | Hoorn Study 2 | 2483 | 165 (6.7%) | 0.75 (0.74; 0.75) | 1.48 (1.20; 1.77) | 0.60 (0.44; 0.76) |
| SCORE2 | 8 | Rotterdam Study | 3282 | 158 (4.8%) | 0.75 (0.75; 0.75) | 1.38 (1.13; 1.62) | 0.00 (-0.16; 0.16) |

n.b. External validation of three guideline recommended prediction models, such as PCE, SCORE2, and QRISK3 for assessing the 10-year risk of major CVD. Point estimates are presented alongside 95% CI. Abbreviations: major cardiovascular disease (CVD) - a composite of coronary heart disease, ischemic stroke, peripheral arterial disease, atrial fibrillation, or heart failure; calibration slope (CS), calibration in the large (CIL).

**Table 6:** External validation of three guideline recommended prediction models for the 10-year risk of CVD.

| Model | No. predictors | Study | Total sample size | No. of CVD events (%) | C-statistic | CS | CIL |
|---|---|---|---|---|---|---|---|
| PCE | 9 | UK Biobank | 93676 | 5687 (6.1%) | 0.71 (0.70; 0.72) | 0.60 (0.57; 0.62) | 0.20 (0.17; 0.23) |
| PCE | 9 | SABRE | 3861 | 366 (9.5%) | 0.64 (0.61; 0.67) | 0.36 (0.28; 0.44) | 0.94 (0.82; 1.06) |
| PCE | 9 | Hoorn Study 1 | 2248 | 359 (16.0%) | 0.71 (0.71; 0.72) | 0.68 (0.57; 0.79) | 0.60 (0.48; 0.72) |
| PCE | 9 | Hoorn Study 2 | 2483 | 155 (6.2%) | 0.74 (0.74; 0.74) | 0.70 (0.55; 0.86) | 0.73 (0.56; 0.90) |
| PCE | 9 | Rotterdam Study | 3282 | 118 (3.6%) | 0.76 (0.76; 0.76) | 0.77 (0.63; 0.92) | -0.42 (-0.61; -0.22) |
| QRISK3 | 22 | UK Biobank | 93676 | 5687 (6.1%) | 0.73 (0.72; 0.74) | 0.96 (0.93; 1.00) | -0.56 (-0.59; -0.54) |
| SCORE2 | 8 | UK Biobank | 93676 | 5687 (6.1%) | 0.72 (0.71; 0.73) | 1.21 (1.16; 1.26) | 0.24 (0.21; 0.26) |
| SCORE2 | 8 | SABRE | 3861 | 366 (9.5%) | 0.70 (0.67; 0.74) | 1.23 (1.04; 1.41) | 0.90 (0.79; 1.01) |
| SCORE2 | 8 | Hoorn Study 1 | 2248 | 359 (16.0%) | 0.72 (0.72; 0.72) | 1.39 (1.18; 1.61) | 0.90 (0.78; 1.02) |
| SCORE2 | 8 | Hoorn Study 2 | 2483 | 155 (6.2%) | 0.75 (0.75; 0.75) | 1.50 (1.21; 1.80) | 0.54 (0.37; 0.70) |
| SCORE2 | 8 | Rotterdam Study | 3282 | 118 (3.6%) | 0.77 (0.76; 0.77) | 1.49 (1.21; 1.78) | -0.31 (-0.50; -0.13) |

n.b. External validation of three guideline recommended prediction models, such as PCE, SCORE2, and QRISK3 for assessing the 10-year risk of CVD. Point estimates are presented alongside 95% CI. Abbreviations: cardiovascular disease (CVD) - a composite of coronary heart disease, ischemic stroke, peripheral arterial disease, calibration in the large (CIL), calibration slope (CS).

**Table 7:** The partial model predicting 10-years risk of major CVD with the best and worst discriminative performance.

| Study | Min c-statistic | No. of predictors for the lowest c-statistic | Max c-statistic | No. of predictors for the highest c-statistic |
|---|---|---|---|---|
| UK Biobank | 0.70 (0.70; 0.70) | 2 | 0.73 (0.73; 0.73) | 14 |
| SABRE | 0.63 (0.60; 0.66) | 4 | 0.69 (0.66; 0.72) | 9 |
| Hoorn Study 1 | 0.70 (0.70; 0.71) | 3 | 0.75 (0.75; 0.75) | 8 |
| Hoorn Study 2 | 0.69 (0.69; 0.69) | 3 | 0.74 (0.74; 0.74) | 9 |
| Rotterdam Study | 0.69 (0.69; 0.70) | 4 | 0.75 (0.75; 0.76) | 9 |

n.b. The partial model predicting 10-years risk of major CVD with the best and worst discriminative performance. Point estimates are presented alongside 95% CI. See discrimination and calibration metrics in the Appendix Data 2 - 3.

**Table 8:** The partial model predicting 10-years risk of major CVD with the best and worst calibration metrics.

| Study | Min CS | CIL for min CS | No. of predictors for the min CS | Max CS | CIL for max CS | No. of predictors for the max CS |
|---|---|---|---|---|---|---|
| UK Biobank | 1.09 (1.05; 1.13) | 0.12 (0.10; 0.15) | 4 | 1.14 (1.10; 1.17) | 0.14 (0.11; 0.16) | 13 |
| SABRE | 0.74 (0.57; 0.90) | 0.54 (0.44; 0.65) | 11 | 1.05 (0.88; 1.21) | 0.49 (0.38; 0.60) | 9 |
| Hoorn Study 1 | 1.20 (1.01; 1.39) | 0.72 (0.61; 0.83) | 4 | 1.55 (1.33; 1.76) | 0.23 (0.12; 0.34) | 8 |
| Hoorn Study 2 | 1.08 (0.83; 1.34) | 0.13 (-0.03; 0.29) | 5 | 1.40 (1.12; 1.67) | 0.09 (-0.07; 0.25) | 6 |
| Rotterdam Study | 1.03 (0.80; 1.26) | -0.42 (-0.58; -0.26) | 5 | 1.40 (1.15; 1.65) | -0.48 (-0.64; -0.32) | 7 |

n.b. The partial model predicting 10-years risk of major CVD with the best and worst calibration metrics: calibration slope (CS) and calibration in the large (CIL). Point estimates are presented alongside 95% CI. See discrimination and calibration metrics in the Appendix Data 2 - 3.

**Table 9:** Participant subgroup sizes across five study cohorts.

| Subgroup | UK Biobank | SABRE | Hoorn Study 1 | Hoorn Study 2 | Rotterdam Study |
|---|---|---|---|---|---|
| Age: | | | | | |
| $< 65$ years | 76793 | 3764 | 1485 | 2413 | 3055 |
| $\geq 65$ years | 16883 | 97 | 763 | 70 | 227 |
| Sex: | | | | | |
| Female | 52253 | 967 | 1250 | 1326 | 1932 |
| Male | 41423 | 2894 | 998 | 1157 | 1350 |
| T2DM status: | | | | | |
| without T2DM | 87339 | 3611 | 2085 | 2384 | 3103 |
| with T2DM | 6337 | 250 | 163 | 99 | 179 |
| Ethnicity | European: 88517<br>South Asians: 1083<br>Chinese: 313<br>Asian or Asian British: 696<br>African Caribbean: 1572<br>Mixed: 1495 | European: 1872<br>Asian: 1400<br>African: 589 | Dutch: 2228<br>Other: 20 | Dutch: 2401<br>Other: 82 | European: 3005<br>non-European: 242 |
| Education level: | | | | | |
| Primary | | | 723 | 87 | 884 |
| Secondary | N/A | N/A | 1386 | 1722 | 168 |
| University | | | 133 | 766 | 200 |
| Townsend deprivation index: | | | | | |
| Least deprived | 23486 | 1017 | | | |
| Middle deprived | 46637 | 1882 | N/A | N/A | N/A |
| Most deprived | 23553 | 962 | | | |

**Table 10:** Number and proportion of major CVD events in each participant subgroup across the five study cohorts.

| Subgroup | UK Biobank | SABRE | Hoorn Study 1 | Hoorn Study 2 | Rotterdam Study |
|---|---|---|---|---|---|
| Age: | | | | | |
| $<$ 65 years | 5196 (6.8) | 372 (9.9) | 186 (12.5) | 151 (6.3) | 125 (4.1) |
| $\geq$ 65 years | 2698 (16.0) | 16 (16.5) | 222 (29.1) | 14 (20.) | 33 (14.5) |
| Sex: | | | | | |
| Female | 3216 (6.2) | 66 (6.0) | 175 (14.0) | 58 (4.4) | 61 (3.2) |
| Male | 4678 (11.3) | 322 (11.1) | 233 (23.2) | 107 (9.2) | 97 (7.2) |
| T2DM status: | | | | | |
| without T2DM | 6619 (7.6) | 322 (8.9) | 350 (16.8) | 145 (6.1) | 3103137 (4.4) |
| with T2DM | 1275 (20.1) | 66 (26.4) | 58 (35.6) | 20 (20.2) | 21 (11.7) |
| Ethnicity | European: 7521 (8.5)<br>South Asians: 97 (9.0)<br>Chinese: 13 (4.2)<br>Asian or Asian British: 77 (11.1)<br>African Caribbean: 88 (5.6)<br>Mixed: 98 (6.6) | European: 165 (8.8)<br>Asian: 196 (14.0)<br>African: 27 (4.6) | Dutch: 407 (18.3)<br>Other: 1 (5.0) | Dutch: 161 (6.7)<br>Other: 4 (4.9) | European: 147 (4.9)<br>non-European: 9.0 (3.7) |
| Education level: | | | | | |
| Primary | | | 158 (21.9) | 8 (9.2) | 58 (6.6) |
| Secondary | N/A | N/A | 227 (16.4) | 115 (6.7) | 92 (4.2) |
| University | | | 21 (15.8) | 40 (5.2) | 8 (4.0) |
| Townsend deprivation index: | | | | | |
| Least deprived | 1847 (7.9) | 84 (8.3) | | | |
| Middle deprived | 3799 (8.1) | 209 (11.1) | N/A | N/A | N/A |
| Most deprived | 2248 (9.5) | 95 (9.9) | | | |

**Table 11:** Comparison of C-statistic quartiles across subsets of partial models predicting 10-year risk of major CVD.

| | UKB | SABRE | HS1 | HS2 | RS |
|---|---|---|---|---|---|
| Models without lipids and/or BP Q1 C-statistic | 0.71 | 0.66 | 0.72 | 0.71 | 0.71 |
| Models without lipids and/or BP Q2 C-statistic | 0.72 | 0.67 | 0.72 | 0.71 | 0.72 |
| Models without lipids and/or BP Q3 C-statistic | 0.72 | 0.67 | 0.73 | 0.72 | 0.73 |
| Models with lipids and/or BP Q1 C-statistic | 0.71 | 0.67 | 0.73 | 0.71 | 0.72 |
| Models with lipids and/or BP Q2 C-statistic | 0.72 | 0.67 | 0.73 | 0.72 | 0.73 |
| Models with lipids and/or BP Q3 C-statistic | 0.72 | 0.68 | 0.74 | 0.73 | 0.74 |

n.b. Abbreviations: UK Biobank (UKB), Southall and Brent Revisited (SABRE), Hoorn Study 1 (HS1), Hoorn Study 2 (HS2), Rotterdam Study (RS), Blood Pressure (BP), quartile 1 (Q1), quartile 2 (Q2, median), quartile 3 (Q3).

#### 1.3 Figures

**Figure 1:** Discriminative performance of partial models predicting 10-Year major CVD or CVD.

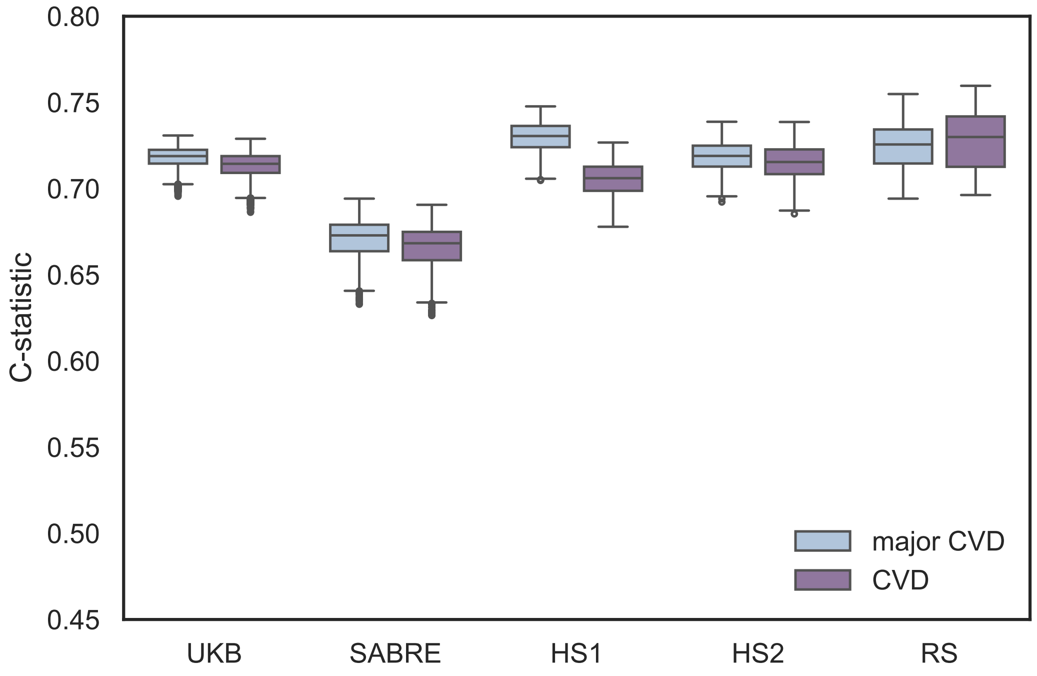

n.b. Cardiovascular disease (CVD) is defined as a composite of coronary heart disease, ischemic stroke, peripheral arterial disease. Major cardiovascular disease (CVD) is an extended definition of CVD that additionally includes atrial fibrillation and heart failure. Abbreviations: UK Biobank (UKB), Southall and Brent Revisited (SABRE), Hoorn Study 1 (HS1), Hoorn Study 2 (HS2), Rotterdam Study (RS).

**Figure 2:** Comparing the performance of guideline recommend models for CVD on their ability to predict CVD and major CVD at 10 years.

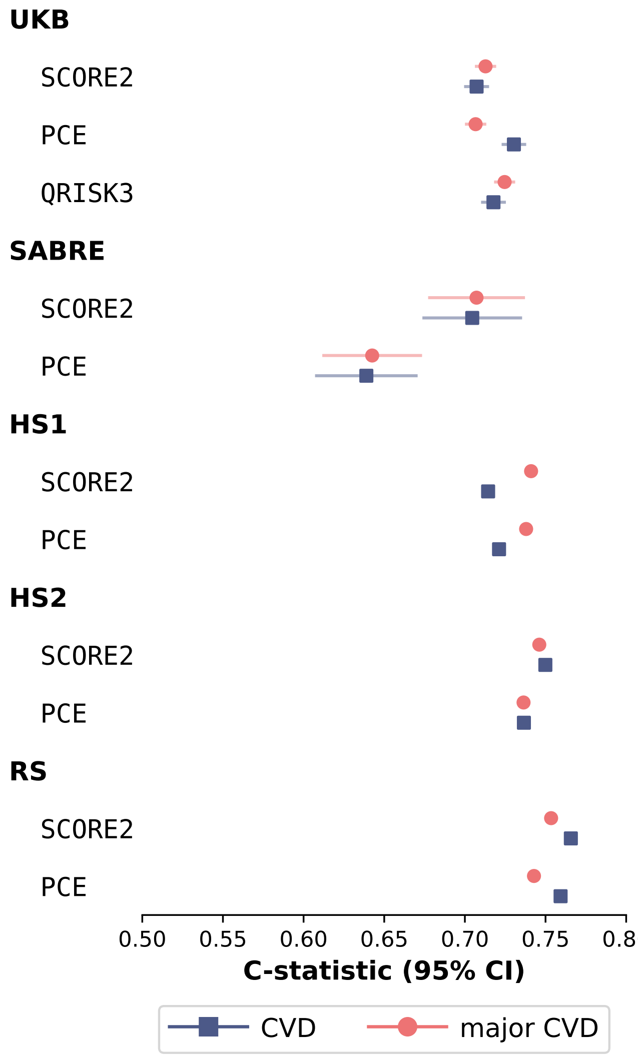

n.b. n.b. Cardiovascular disease (CVD) is defined as a composite of coronary heart disease, ischemic stroke, peripheral arterial disease. Major cardiovascular disease (CVD) is an extended definition of CVD that additionally includes atrial fibrillation and heart failure. Abbreviations: UK Biobank (UKB), Southall and Brent Revisited (SABRE), Hoorn Study 1 (HS1), Hoorn Study 2 (HS2), Rotterdam Study (RS), Systematic Coronary Risk Evaluation 2 (SCORE2) [1], Pooled Cohort Equations (PCE) [2], QRISK3 [3].

**Figure 3:** The calibration slope of the set of partial models predicting 10-year risk of major CVD across five cohort studies.

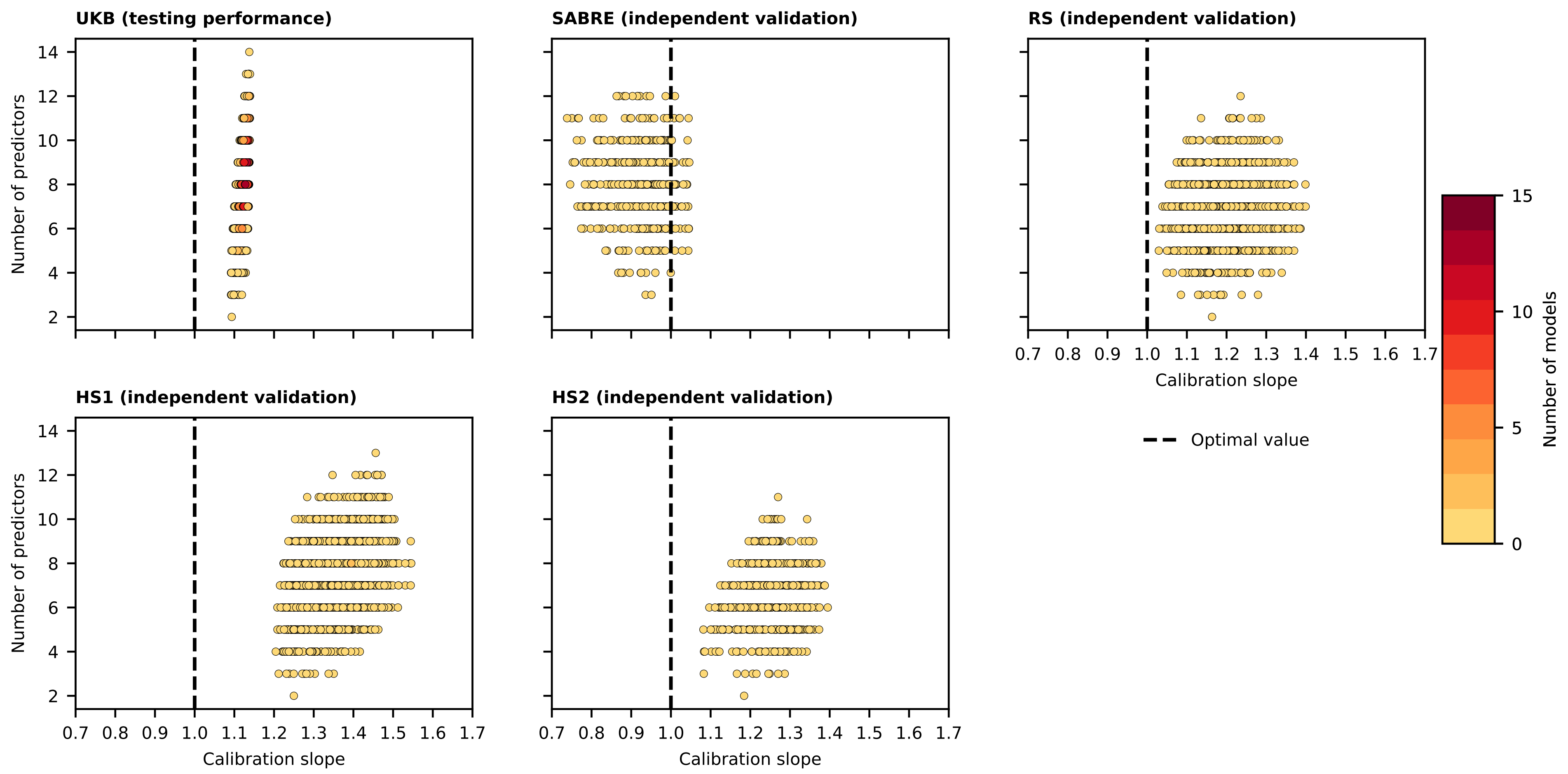

n.b. The calibration slope was calculated for both the set of partial models and established models (PCE, SCORE2, and QRISK3) across five cohort studies: UK Biobank (UKB) (93,676 participants, with 7,894 (8.4%) experiencing a major CVD event over 10 years), Southall and Brent Revisited (SABRE) (3,861 participants, 388 (10.1%) events), the Hoorn Study 1 (HS1) (2,248 participants, 359 (16.0%) events), the Hoorn Study 2 (HS2) (2,483 participants, 165 (6.7%) events, and the Rotterdam Study (RS) (8,882 participants, 1,014 (11.4%) events). The calibration slopes (CS) (x-axis) are plotted against the number of predictors considered by the major CVD risk prediction models (y-axis). The PCE CS was 0.60 in UKB, 0.36 in SABRE, 0.78 in

**Figure 4:** The calibration in the large of the set of partial models predicting 10-year risk of major CVD across five cohort studies.

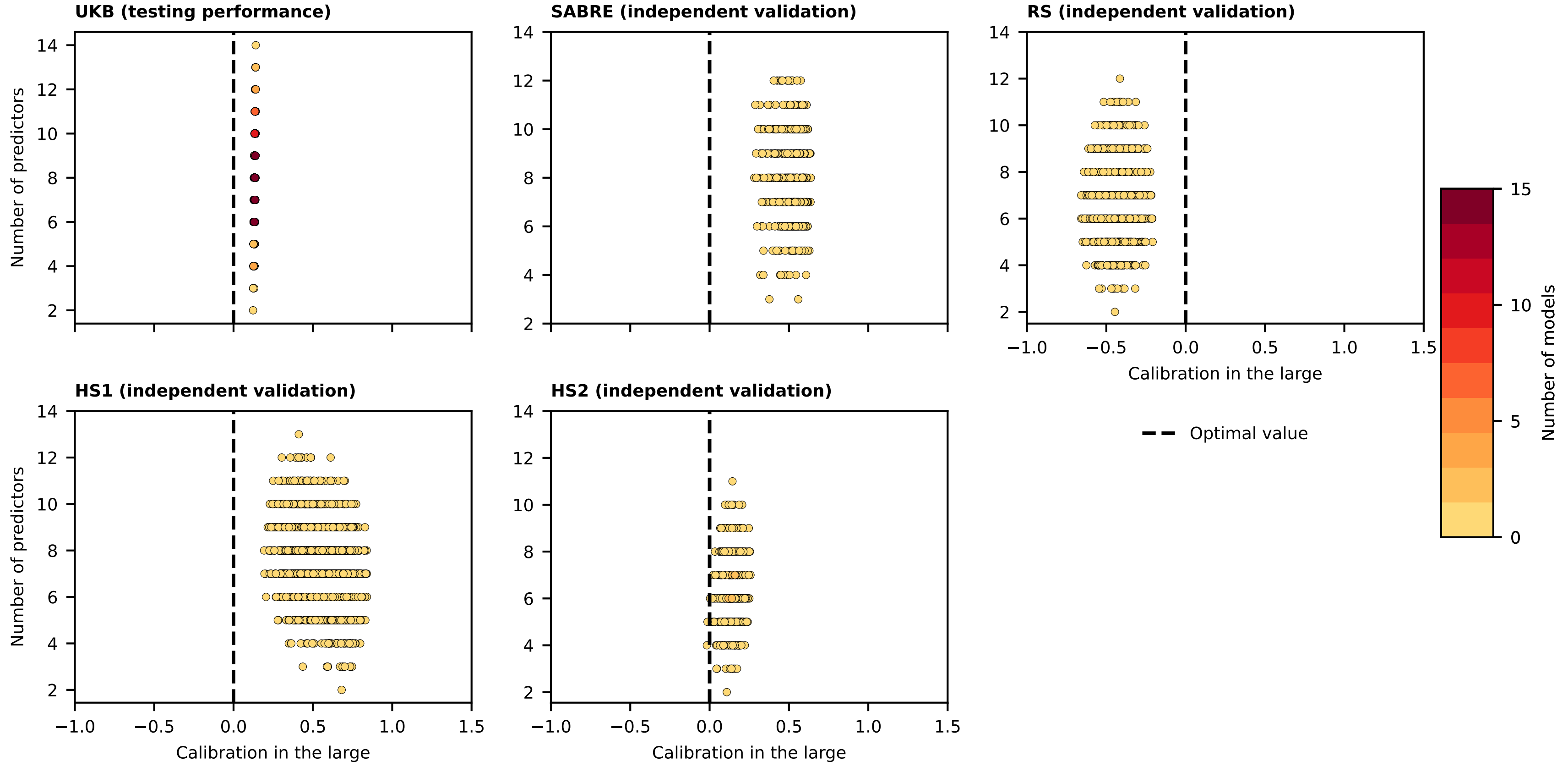

n.b. The calibration in the large was calculated for both the set of partial models and established models (PCE, SCORE2, and QRISK3) across five cohort studies: UK Biobank (UKB) (93,676 participants, with 7,894 (8.4%) experiencing a major CVD event over 10 years), Southall and Brent Revisited (SABRE) (3,861 participants, 388 (10.1%) events), the Hoorn Study 1 (HS1) (2,248 participants, 359 (16.0%) events), the Hoorn Study 2 (HS2) (2,483 participants, 165 (6.7%) events), and the Rotterdam Study (RS) (8,882 participants, 1,014 (11.4%) events). The calibration in the large (x-axis) is plotted against the number of predictors considered by the major CVD risk prediction models (y-axis). The PCE calibration in the large (CIL) was 0.59 in UKB, 1.02 in SABRE, 0.79 in the HS1, 0.80 in HS2, and -0.25 in RS. The QRISK3 CIL was -0.19 in UKB. The SCORE2 CIL was 0.6 in UKB, 0.97 in SABRE, 1.06 in HS1, 0.6 in HS2, and 0.0 in RS. See calibration metrics in the Appendix Data 2.

**Figure 5:** The median age and sex feature importance of the set of partial models predicting 10-year risk of major CVD assessed across five study cohorts.

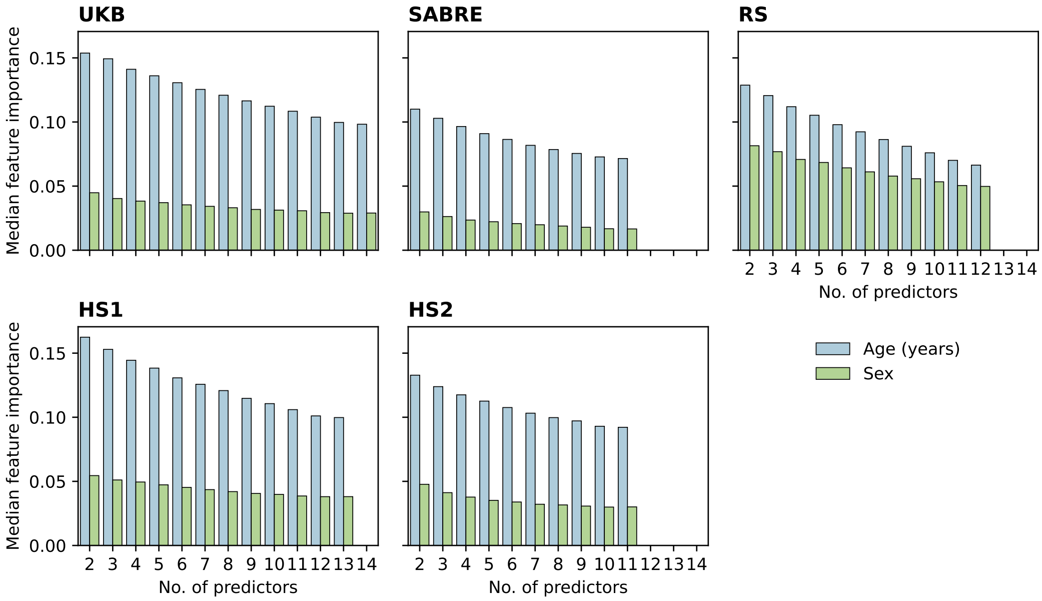

n.b. The median age and sex feature importance of the set of partial models predicting 10-year risk of major CVD assessed across five study cohorts. The permutation feature importance assesses the c-statistic change in study cohort; iteratively the values of each predictor were randomly assigned to an individual after which the c-statistic was re-estimated with these permuted data and the difference in performance used as an estimate of predictor contribution to the model's predictive potential. See feature importance in the Appendix Data 3. Abbreviations: UK Biobank (UKB), Southall and Brent Revisited (SABRE), Rotterdam Study (RS), Hoorn Study 1 (HS1), Hoorn Study 2 (HS2).

**Figure 6:** Discriminative performance of the set of partial models predicting 10-year risk of major CVD per UK Biobank participant subgroup.

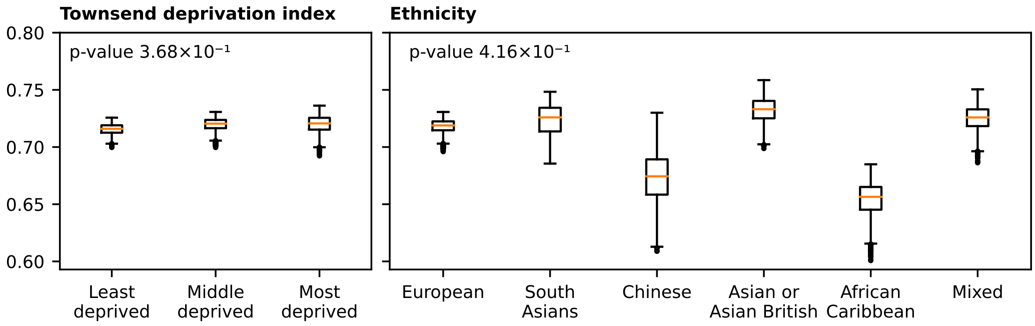

n.b. Discriminative performance of the set of partial models predicting 10-year risk of major CVD per UK Biobank participant subgroup. The p-value is calculated using the Wilcoxon test for subgroups with two categories and the Kruskal-Wallis test for subgroups with more than two categories. See participant subgroup sizes in Appendix Table 10 and subgroup definitions in the Appendix 2.

**Figure 7:** Discriminative performance of the set of partial models predicting 10-year risk of major CVD per SABRE participant subgroup.

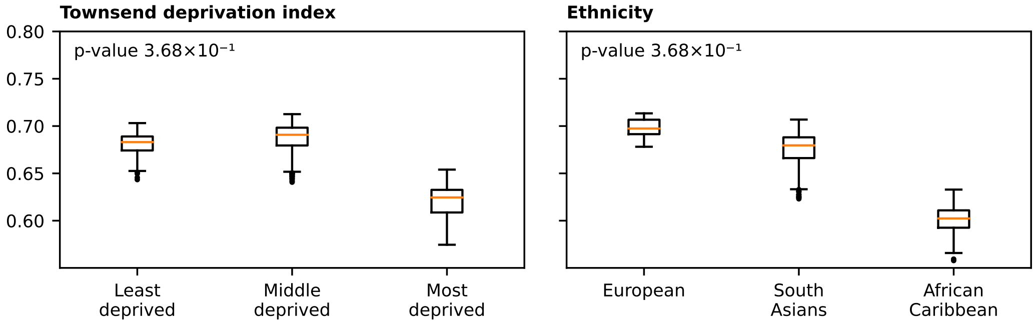

n.b. Discriminative performance of the set of partial models predicting 10-year risk of major CVD per SABRE participant subgroup. The p-value is calculated using the Wilcoxon test for subgroups with two categories and the Kruskal-Wallis test for subgroups with more than two categories. Refer to Appendix Table 10 for participant subgroup sizes and Appendix 2 for subgroup definitions.

**Figure 8:** Discriminative performance of the set of partial models predicting 10-year risk of major CVD per Hoorn Study 1 participant subgroup.

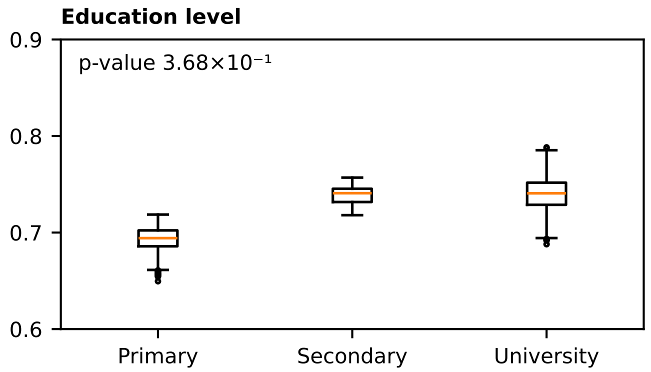

n.b. Discriminative performance of the set of partial models predicting 10-year risk of major CVD per Hoorn Study 1 participant subgroup. The p-value is calculated using the Wilcoxon test for subgroups with two categories and the Kruskal-Wallis test for subgroups with more than two categories. Refer to Appendix Table 10 for participant subgroup sizes and Appendix 2 for subgroup definitions.

**Figure 9:** Discriminative performance of the set of partial models predicting 10-year risk of major CVD per Hoorn Study 2 participant subgroup.

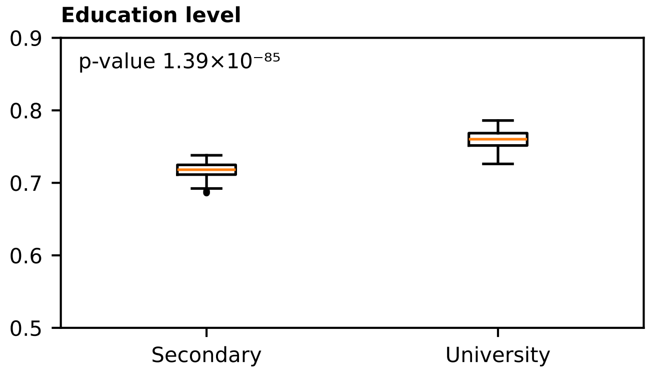

n.b. Discriminative performance of the set of partial models predicting 10-year risk of major CVD per Hoorn Study 2 participant subgroup. The p-value is calculated using the Wilcoxon test for subgroups with two categories and the Kruskal-Wallis test for subgroups with more than two categories. Refer to Appendix Table 10 for participant subgroup sizes and Appendix 2 for subgroup definitions.

**Figure 10:** Discriminative performance of the set of partial models predicting 10-year risk of major CVD per Rotterdam Study participant subgroup.

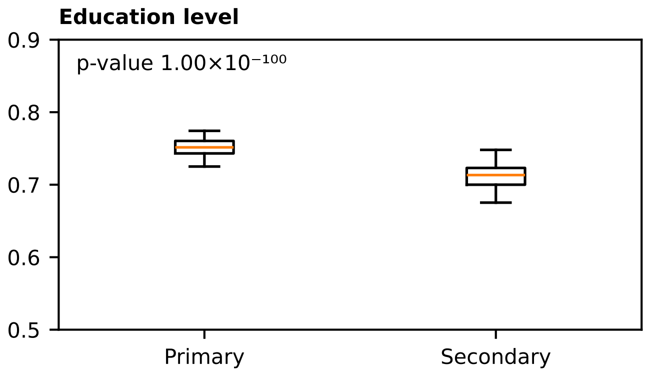

n.b. Discriminative performance of the set of partial models predicting 10-year risk of major CVD per Rotterdam Study participant subgroup. The p-value is calculated using the Wilcoxon test for subgroups with two categories and the Kruskal-Wallis test for subgroups with more than two categories. Refer to Appendix Table 10 for participant subgroup sizes and Appendix 2 for subgroup definitions.
